# Local and Distant responses to single pulse electrical stimulation reflect different forms of connectivity

**DOI:** 10.1101/2020.10.13.20212266

**Authors:** Britni Crocker, Lauren Ostrowski, Ziv M. Williams, Darin D. Dougherty, Emad N. Eskandar, Alik S. Widge, Catherine J. Chu, Sydney S. Cash, Angelique C. Paulk

## Abstract

**Background:** Measuring connectivity in the human brain can involve innumerable approaches using both noninvasive (fMRI, EEG) and invasive (intracranial EEG or iEEG) recording modalities, including the use of external probing stimuli, such as direct electrical stimulation.

**Objective/Hypothesis:** To examine how different measures of connectivity correlate with one another, we compared ‘passive’ measures of connectivity during resting state conditions map to the more ‘active’ probing measures of connectivity with single pulse electrical stimulation (SPES).

**Methods:** We measured the network engagement and spread of the cortico-cortico evoked potential (CCEP) induced by SPES at 53 total sites across the brain, including cortical and subcortical regions, in patients with intractable epilepsy (N=11) who were undergoing intracranial recordings as a part of their clinical care for identifying seizure onset zones. We compared the CCEP network to functional, effective, and structural measures of connectivity during a resting state in each patient. Functional and effective connectivity measures included correlation or Granger causality measures applied to stereoEEG (sEEGs) recordings. Structural connectivity was derived from diffusion tensor imaging (DTI) acquired before intracranial electrode implant and monitoring (N=8).

**Results:** The CCEP network was most similar to the resting state voltage correlation network in channels near to the stimulation location. In contrast, the distant CCEP network was most similar to the DTI network. Other connectivity measures were not as similar to the CCEP network.

**Conclusions:** These results demonstrate that different connectivity measures, including those derived from active stimulation-based probing, measure different, complementary aspects of regional interrelationships in the brain.

## Introduction

Brain connectivity measures are a popular approach to quantify brain networks and the inter-regional coupling between brain areas. Typically, brain network estimates are categorized into three distinct types: functional, effective, and structural [1]. Functional connectivity identifies channels or brain regions with correlated activity, and is measured in the neural signal, such as in the local field potential (LFP), using methods such as correlation, cross-correlation, and magnitude-squared coherence [2]. Effective connectivity identifies whether channels or brain regions may have causal influence on one another and includes methods such as weighted phase lag index and Granger causality [1]. Diffusion tensor imaging (DTI) allows for estimates of structural connectivity by measuring the anisotropy of water molecules, allowing for a partial reconstruction of the white matter tracts in the brain [3]. All of these measures of connectivity can occur without external stimuli or perturbations to the system.

In contrast, connectivity measures derived from cortico-cortical evoked potentials (CCEPs) involve external perturbations to the system in which evoked potentials are elicited by direct intracranial single pulse electrical stimulation [4,5]. CCEPs can be used to estimate the connectivity of multiple different cortical areas [4–13]. CCEP connectivity is dynamic and can reveal changes related to state of consciousness [14] or underlying pathology [13,15,16]. Importantly, CCEPs are not symmetric in their waveform nor spatial spread and can therefore be used to create directed network graphs [17–19]. Thus, CCEPs offer many advantages over available tools for estimating brain networks. In a recent study using data from hundreds of individuals, researchers found there was a significant correlation between distance from the recording site and both the latency of the CCEP waveform and identified probability connectivity networks [9].

Within the theoretical framework of brain network categorization, it has been hypothesized that CCEP networks represent functional or effective connectivity [4,9,20]. CCEP networks do correspond with other functional connectivity measures, such as resting state high gamma coherence on electrocorticography (ECoG; [19]) and resting state functional magnetic resonance imaging (fMRI; [21]). Diffusion tractography has been shown to correlate with CCEP networks in the language system [7] and has been used to predict changes in post-stimulus brain activity [22]. Here, we test the similarities between CCEP connectivity estimates and six common estimates of brain connectivity, including 3 measures of functional connectivity (correlation, cross-correlation, magnitude square coherence), 2 measures of effective connectivity (Granger causality, phase lag index) and a robust measure of structural connectivity (probabilistic tractography). We find a significant similarity between CCEPs and all coupling measures that persists even after distance correction, suggesting the CCEPs are a reliable measure of brain connectivity. However, the degree to which the measures correlate with the CCEP is dependent on distance and type of connectivity measure.

## Methods

### Data Collection

Neurophysiological data was recorded from 11 patients (6 females and 5 males) of ages 19 to 57 (mean 36.9, std 12.5) undergoing invasive monitoring for intractable epilepsy using bilateral intracranial depth electrodes. Depth electrodes (Ad-tech Medical, Racine WI, USA, or PMT, Chanhassen, MN, USA) had diameters of 0.8–1.0 mm and consisted of 8-16 platinum/iridium-contacts 1-2.4 mm long. The placement of these electrodes was determined solely by clinical criteria, with most electrodes in the frontal or temporal lobe. All patients provided informed consent to participate in any experiments, which were approved by the Massachusetts General Hospital Institutional Review Board and the Massachusetts Institute of Technology Committee On the Use of Humans as Experimental Subjects following the National Institutes of Health and Army Human Research Protection Office guidelines.

Voltage data from implanted electrodes were recorded at a sampling rate of 2000 Hz using a Neural Signal Processor system (Blackrock, Inc.). In addition to these electrophysiological recordings, each patient underwent a T1-weighted structural magnetic resonance imaging (MRI) scan which was coregistered to the post-operative computed tomography (CT) scan to identify electrode locations [23,24]. Distance between channels is defined as the Euclidean distance between the average locations for each bipolar electrode pair.

### Stimulation

To evoke cortico-cortical evoked potentials (CCEPs), biphasic bipolar single pulses of stimulation were applied across pairs of adjacent electrodes with a 90us pulse width and a 53us inter-pulse interval at 6 or 7 mA (**Figure 1A**). This stimulus strength was chosen as one that has been previously reported to evoke CCEPs while remaining low enough to avoid afterdischarges [17]. These pulses were delivered by the Blackrock CereStim stimulator at 3-second intervals with +/- 500 ms random jitter with uniform distribution and at an amplitude below the threshold, above which stimulation would induce afterdischarges or for any perceptual or behavior effects reported by the patients. In one participant, these single pulses were delivered interspersed between other trains of stimulation. The number of trials ranged from 20 to 337 (mean 47), and stimulation occurred in one to nine sites per participant. The stimulation sites were not in the seizure onset zone or areas where early seizure activity spreads for any patient, and all stimulation experiments were performed while the patients were awake, after the patients resumed antiepileptic medication, and were under medical supervision.

**Figure 1:**
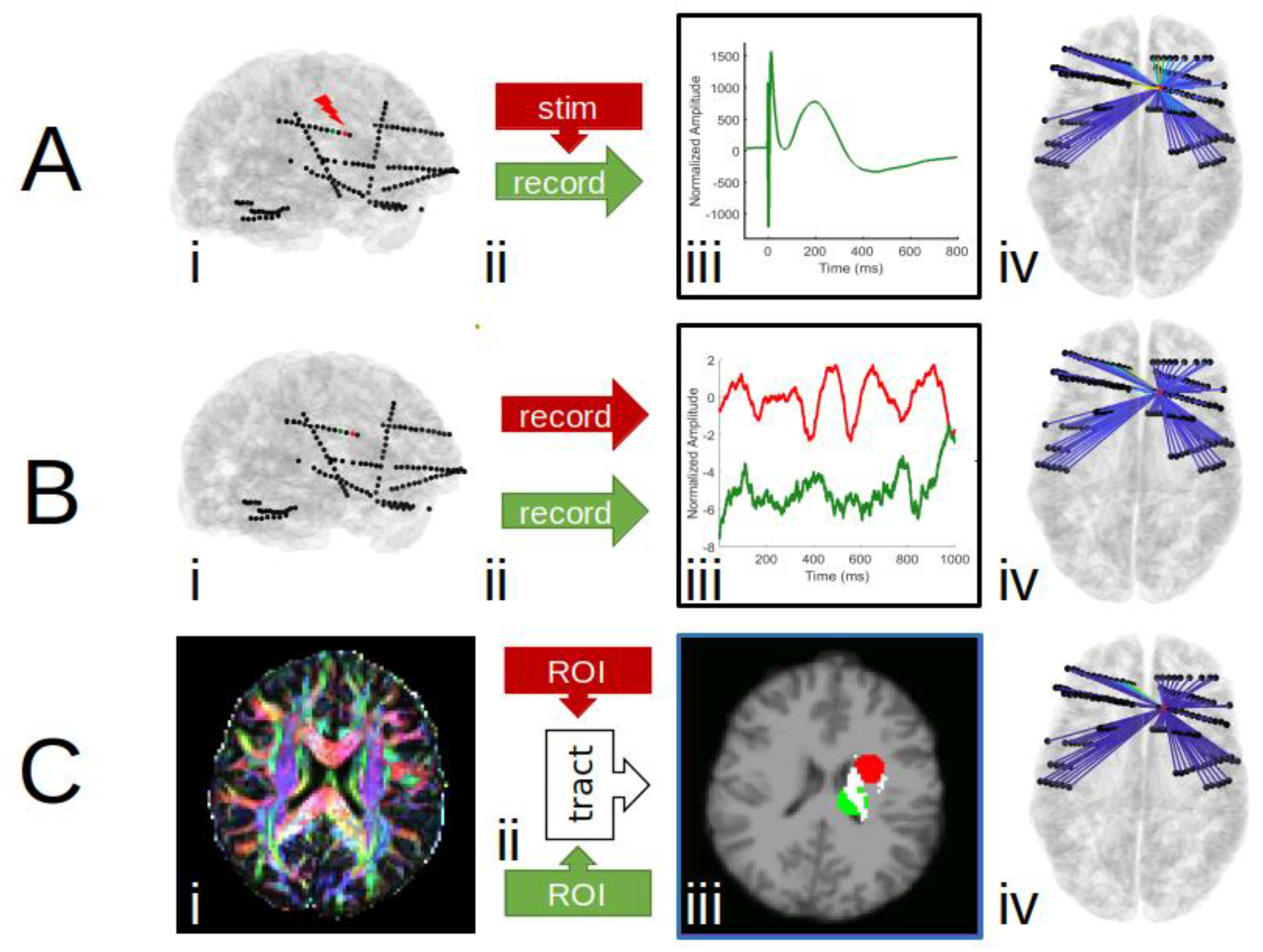
an overview of how networks are computed and compared. First Column (i). reconstruction in one representative participant of data collection by stimulating one channel and recording the others (A), recording neurophysiological signals at rest in all channels (B), or collecting diffusion tensor imaging (C). In the first two panels of column i, channel locations are shown in the 3-D reconstruction of the pial surface from structural MRI, with the stimulation channel marked in red and one of the recording channels marked in green. **B**.Second Column (ii) schematic representations of the three data collection methods. For CCEP networks, one channel is stimulated and the others are recorded. For functional and effective networks, all channels are recorded. For structural networks, the channel locations are used to create regions of interest (ROIs) for both seed and target masks; tractography is then computed between pairs of ROIs. **C**.Third Column (iii) representative data samples for each type of data collection method. An example CCEP is shown in row A. Resting state electrophysiology is shown in both the stimulation channel (red) and a non-stimulation channel (green) before the stimulation experiment in row B. In row C, the ROI created from the stimulation channel location (red) and a non-stimulation channel (green) is shown overlaid on the structural MRI. The tract between the two ROIs is highlighted in white. **D**. example networks. Last Column (iv) from each network construction method overlaid on representative anatomy. Bluer lines represent weaker pairwise connections between the stimulation channel or ROI and all other channels or ROIs, while yellower lines represent stronger connections. Similarity scores are then calculated as the correlation between a given network, transformed into a vector, and the CCEP network, also transformed into a vector.

### CCEP Networks

To estimate CCEP networks, recordings were first analyzed using custom software in MATLAB (Mathworks; Natick, MA). The data were re-referenced using a bipolar reference scheme between neighboring electrodes then divided into 2-second epochs time-locked to the delivery of the stimulus, including one second pre- and one second post-stimulation. Any peri-stimulus data within 3 ms of the stimulation pulse were excluded from analysis, before and after the stimulus. All channels and trials were visually inspected for artifacts, and any channels or trials with large artifacts were excluded from analysis. In the one second following stimulation, the voltage recordings for each channel were averaged across trials and then normalized by the standard deviation of the voltage in the one second before stimulation (**Figure 1Aiii**). CCEP responses were quantified per bipolar recording by the maximum absolute amplitude of the average evoked potential in this normalized signal [21].

These CCEP responses constitute a one-dimensional CCEP network representing the connectivity between the stimulation site and each recording channel. Stimulation at some sites failed to produce a response in any recording channels. To avoid constructing or comparing empty CCEP networks, stimulation sites were only included in analysis if CCEPs greater than 6 standard deviations above baseline could be measured in any recording channels. Since CCEP responses can be asymmetric, these one-dimensional CCEP networks are to be understood as directed networks from the stimulation site to the recording sites (**Figure 1A**).

We then compared the CCEP network to other connectivity metrics (see Network Comparisons section). Two networks could share many connections between nodes and still be dissimilar due to a mismatch in the density of connections. To help distinguish this case from completely dissimilar networks, we compared the average network strength between channels that had either “above-threshold” or “below-threshold” CCEP responses. A threshold of 6 standard deviations above baseline was chosen to separate above-threshold and below-threshold CCEP responses measured in the method described above [17,21].

### Functional Connectivity Calculations

For functional connectivity estimates, 5 minutes of recordings while the participants were awake before or after single pulse stimulation sessions were used (**Figure 1A-B**). These data were re-referenced with the same bipolar reference scheme, then bandpass filtered from 1-150 Hz and notch filtered from 58-62 Hz and 118-122 Hz (Butterworth 3rd order, forward and reverse). The recordings were then split contiguously into 1-second epochs. As above, channels and trials identified by visual inspection as having large artifacts were excluded from further analysis.

Correlation and cross-correlation measures were used as estimates of functional connectivity in the time domain. For each 1-second epoch, the correlation and maximum cross-correlation with lags between +/- 250 ms were calculated between the stimulation electrode pairs and all other electrode pairs. For each channel pair, the above-chance threshold of connectivity was determined using a bootstrap method: independently shuffling epochs (with replacement) 10,000 times to destroy the temporal relationship between epochs, then choosing the values at the 2.5^th^ and 97.5^th^ percentile. Then the final network was determined as the percentage of times each correlation or cross-correlation rose above this random chance threshold, yielding values between 0 and 1 [25,26].

Similarly, magnitude-squared coherence was used as an estimate of functional connectivity in the frequency domain. For each 1-second epoch, magnitude-squared coherence was calculated for each frequency from 1 to 150 Hz between stimulation electrode pairs and all other electrode pairs. Again, for each epoch, a 5% threshold was calculated using the above bootstrap method, and the final network for each frequency was given by the percentage of times each MS-coherence estimate rose above the 5% threshold, yielding values between 0 and 1.

### Effective Connectivity Calculations

For effective connectivity estimates, the same 5 minutes of recording were used with the same bipolar reference scheme as for functional connectivity. For pairwise Granger causality estimates, these recordings were split contiguously into 1-second epochs and then decimated from 2000 Hz to 250 Hz in order to reduce computational time. Pairwise Granger causality was estimated using the multivariate Granger causality toolbox [27] with a model order of 10, between every recording channel and the stimulation channel for each stimulation site. Pairwise Granger causality networks are directed networks and were therefore computed in both directions: from each stimulation site to every recording site, and from every recording site to each stimulation site. Weighted phase lag indices were calculated from 4 to 50 Hz with the FieldTrip toolbox using 2-second contiguous epochs in order to capture lower frequency networks [28,29].

### Structural Connectivity Calculations

Diffusion tensor imaging was acquired heterogeneously across 8 of 11 participants included in this data set, with b-values of 700 (1 participant), 1000 (3 participants), and 2000 (4 participants) and number of directions ranging from 28 to 74. Structural connectivity between two regions was determined using the FDT toolbox [30,31]. First, we used BEDPOSTx (Bayesian Estimation of Diffusion Parameters Obtained using Sampling Techniques for modelling Crossing Fibres) to locally model the diffusion parameters (**Figure 1A-C**). The average coordinates of the bipolar electrode pairs from the structural imaging were transformed into diffusion-space using the transformation matrix obtained by coregistration of the structural MRI and the diffusion MRI with FLIRT [32–34]. These transformed coordinates were then used to generate binary masks of spheres of 1-centimeter radius centered on each channel [35]. For each stimulation site, probtrackx2 was then used to calculate the number of streamlines (inferred fiber tracts) from the seed mask to the target mask, with the target mask as both a waypoint and a termination mask (**Figure 1C**). This process was repeated for the stimulation mask as seed mask and recording mask as target mask (generating streamlines from the stimulation channel to the recording channel) and for the stimulation mask as target mask and the recording mask as seed mask (generating streamlines from the recording channel to the stimulation channel). The number of streamlines was then normalized by the total mask volume in order to generate a value for structural connectivity.

### Network Comparisons

Every set of connectivity estimates generated for each stimulation site could be represented as a row in the total weighted adjacency matrix of the complete connectivity matrix across channels for each participant (**Figure 1iv**). Since participants had an unequal number of stimulation sites, we computed the similarity of connectivity estimates for each row (i.e. each stimulation site). To compare CCEP networks to other network estimates, we calculated the similarity score between two networks as the Pearson’s correlation between the two vectors. Unless otherwise noted, all significance calculations are uncorrected t-tests of the Fisher-transformed correlation coefficients – two-sided when comparing two methods, or one-sided when comparing to zero. Confidence intervals were determined using bootstrap-resampling over 1000 iterations.

Distance is a strong confound when comparing any brain connectivity networks, since areas close together are much more likely to be connected [26]. When controlling for distance, we used a partial correlation to compute similarity between two networks. The least squares linear regression is computed between each 1D network and the reciprocal of distance squared; the distance-controlled similarity between the two networks is then the correlation of the residuals from each of these regressions.

## Results

We recorded corticocortical evoked potentials (CCEPs) induced by single pulse intracranial direct electrical stimulation at 53 total sites across 11 participants with intractable epilepsy implanted with stereo-encephalography electrodes for the purposes of monitoring neural activity as part of their clinical care. Sites were largely in prefrontal and temporal cortices. The spread of the CCEP across recording sites was quantified (see **Methods**) and arranged into one-dimensional CCEP networks (**Figure 1Aiv**). To determine whether these CCEP networks reliably reflect functional, effective, or structural connectivity, we then compared the voltage response spread of CCEPs to a battery of measures which reflect each of these three types of connectivity (see **Methods**). For this reason, in addition to voltage and spectral connectivity measures, we compared the stimulation responses to preoperative diffusion tensor imaging (DTI) to perform a partial reconstruction of the white matter tracts in a subset of participants (N=8).

### Functional Connectivity

#### Correlation networks

Correlation-derived connectivity patterns from neural activity during rest (outside of periods of stimulation) shared substantial similarities to the CCEP network. Across the 53 stimulation sites, correlation networks corresponded to CCEP networks with an average similarity score of 0.50 ± 0.24 std (**Figure 2A**). This relationship was reflected in the data; channels with an above-threshold response to stimulation had an average absolute correlation of 0.20 ± 0.18 std to the stimulation channel at rest, compared to 0.037 ± 0.045 std in channels with no response to stimulation (**Figure 2B**). The relatively high variance of correlation among stimulation-responsive channels suggests that correlation may not be useful for predicting whether there would be a CCEP or not, but the large difference between above-threshold and below-threshold channels suggests that correlation is reasonably specific to whether there was an above-threshold response at all.

**Figure 2:**
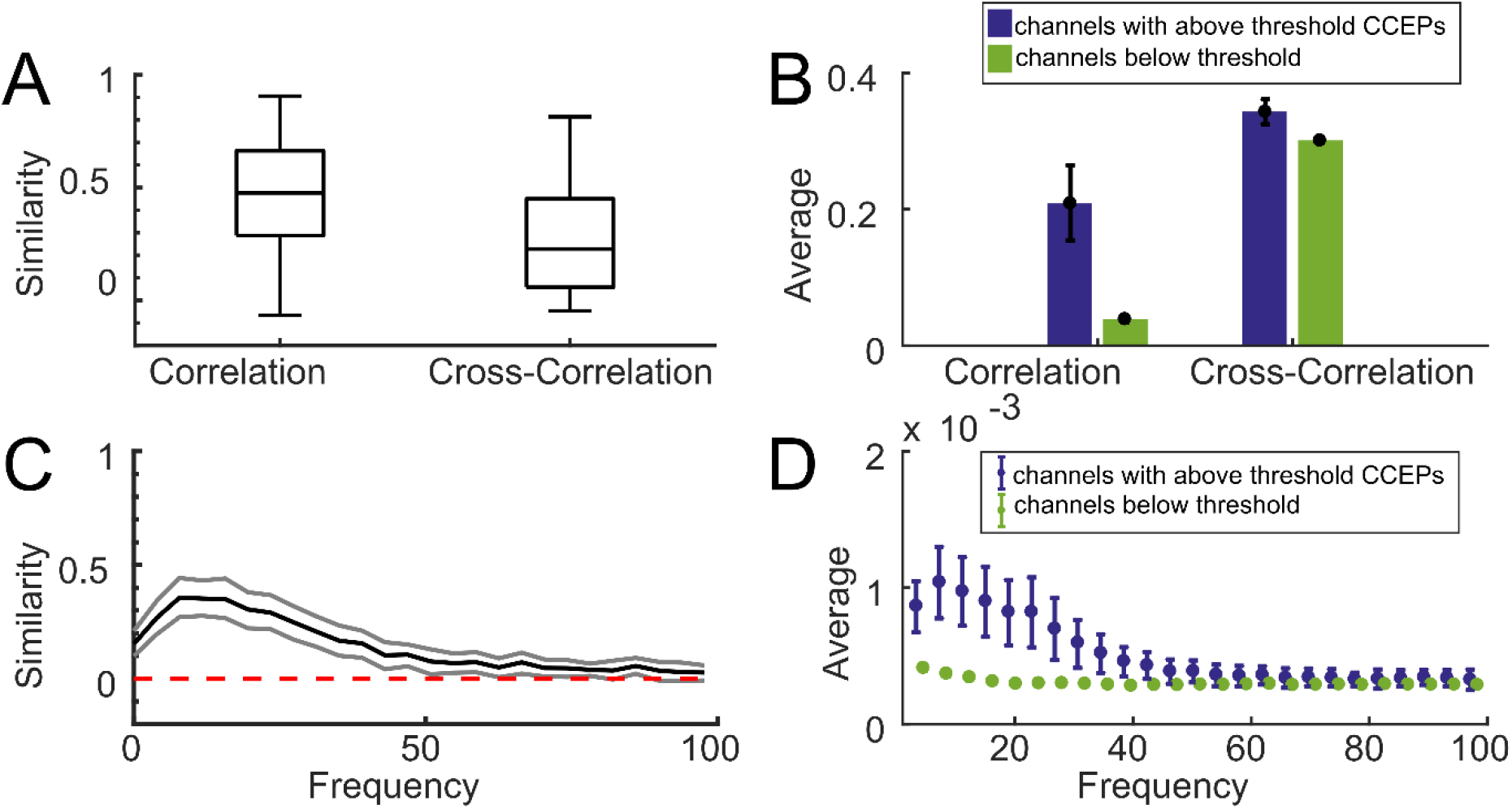
Functional connectivity networks show similarity to stimulation-evoked networks. **A**. Boxplots showing the medians and confidence bounds of the similarity scores between correlation or cross-correlation networks and CCEPs for different stimulation sites. **B**. the average correlation and normalized cross-correlation values for all channels with (blue) or without (green) an above-threshold CCEP. Uncorrected confidence intervals (black error bars) were determined by naive bootstrap resampling. Error bars reflect 95% confidence intervals. **C**. the mean (black) similarity across the 34 stimulation sites for magnitude-squared coherence networks and CCEP networks at each frequency, and the uncorrected 95% confidence intervals at each frequency (grey) determined by naïve bootstrap resampling. **D**. The average unnormalized magnitude-squared coherence for all channels with (blue) or without (green) an above-threshold CCEP, with the same threshold as in B. Uncorrected confidence intervals (error bars) were determined by naive bootstrap resampling.

### Cross-correlation networks

Cross-correlation-derived connectivity patterns are significantly less similar to the CCEP network than correlation (p = 1.5e-08). Across 53 stimulation sites, cross-correlation networks had an average similarity score of 0.28 ± 0.22 std with CCEP networks (**Figure 2A**). Channels with above-threshold CCEPs had a normalized cross-correlation of 0.34 ± 0.069 std to the stimulation channel at rest, compared to 0.3000 ± 0.041 std in channels which were below threshold (**Figure 2B**). While cross-correlation was on average higher in channels with above-threshold CCEPs compared to correlation, cross-correlation networks still were less similar than correlation networks to CCEP networks. This decrease in correspondence between the cross-correlation networks and CCEP networks was caused by the relatively high cross-correlation values between channels which were below the CCEP threshold (indicating non-CCEP responsive locations) and the stimulation channel.

In addition to examining the cross-correlation relationships, we examined whether there was a consistent relationship between cross correlation lag and the CCEP network. We found a non-significant (p = 0.09, 2-sample F test to compare variances) decrease in lag in channel pairs in the CCEP network versus channels not in the CCEP network (data not shown). This difference completely disappeared when we compared only channels with a reasonably high cross correlation value (>0.2), so this trend could just be due to the association between CCEP magnitude and cross-correlation listed above.

### Coherence networks

The similarity between magnitude-squared-coherence-derived networks and CCEP networks was computed over a range of frequencies, with the best agreement occurring at approximately 8 Hz (**Figure 2C**). At 8 Hz, coherence networks corresponded to CCEP networks with an average similarity score of 0.40 ± 0.25 std (**Figure 2C**), which was significantly better than cross-correlation and worse than correlation (p = 1.4e-04 and 1.2e-05, respectively, t-test). Across all frequencies up to 100 Hz, the lower bound of the uncorrected 95% confidence interval for network similarity was above zero. Like correlation, channels with above-threshold CCEPs showed a larger coherence with the stimulation channel than channels with responses below threshold (**Figure 2D**).

### Effective Connectivity

#### Granger causality

Pairwise Granger causality is a directed measure of connectivity, yielding directional estimates of connectivity from the stimulation channel to the recording channel and vice versa. Across the 53 stimulation sites, we calculated the pairwise Granger causality for neural data during rest outside of stimulation and compared this measure to the CCEP networks per stimulation site. Pairwise Granger causality networks from the stimulation channel corresponded to CCEP networks with an average similarity score of 0.38 ± 0.27, while pairwise Granger causality networks from the recording channel corresponded to CCEP networks with an average similarity score of 0.34 ± 0.28 std (**Figure 3A**). No significant difference was found between the two groups (p = 0.39). Channels with above-threshold CCEPs had higher resting state Granger causality values with the stimulation channel at resting state (0.014 ± 0.019 std from the stimulation channel, and 0.017 ± 0.0120 std from the recording channel) than the remaining channels (0.0050 ± 0.012 std from the stimulation channel, and 0.0061 ± 0.014 std from the recording channel; **Figure 3B**). Overall, these similarities are significantly smaller than the functional connectivity correlation similarities (p = 8.0e-4 and 4.1e-04; t-test).

**Figure 3:**
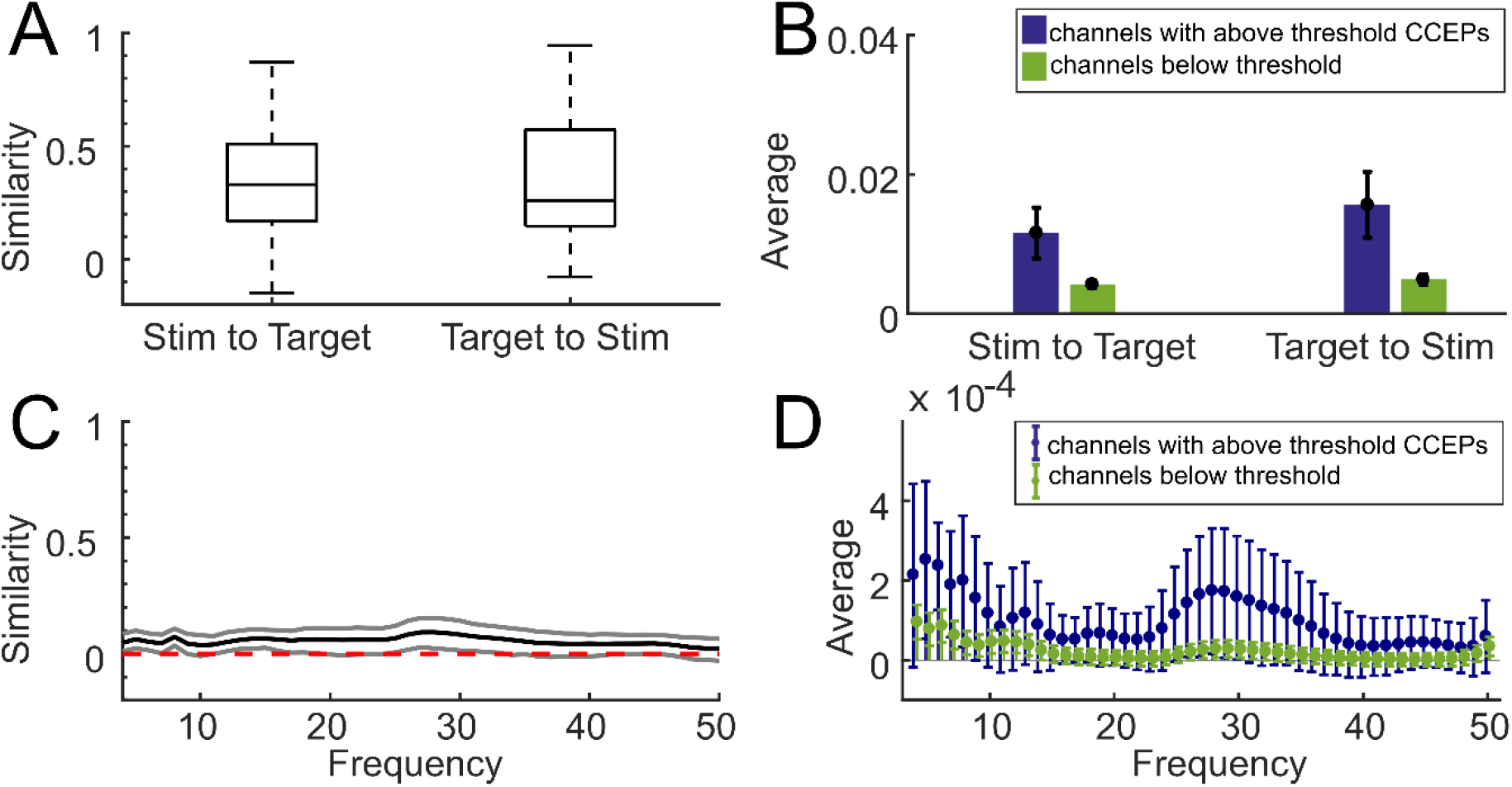
Effective connectivity shows less similarity to stimulation-evoked networks. **A**. boxplots showing the medians and confidence bounds of the similarity scores between pairwise Granger causality networks in both directions and CCEPs across 34 for different stimulation sites. Pairwise Granger causality is a directed measure of connectivity, so calculations from the stimulation site to the recording channel and from the recording channel to the stimulation site are both represented. **B**. The average unnormalized pairwise granger causality values for all channels with (blue) or without (green) an above-threshold CCEP. The presence of a CCEP is defined as any average evoked potential that reaches an absolute value at least 6 standard deviations above baseline mean. Uncorrected confidence intervals (black error bars) were determined by naive bootstrap resampling. Error bars reflect 95% confidence intervals. Every measure of connectivity has its own scale, so absolute values are not as important as the difference between the two groups. **C**. The mean (black) similarity across the 34 stimulation sites for weighted phase lag index networks and CCEP networks at each frequency, and the uncorrected 95% confidence intervals at each frequency (grey) determined by naïve bootstrap resampling. **D**. The average weighted phase lag index for all channels with (blue) or without (green) an above-threshold CCEP, with the same threshold as in **B**. Uncorrected confidence intervals (error bars) were determined by naive bootstrap resampling. Every measure of connectivity has its own scale, so absolute values are not as important as the difference between the two groups.

#### Phase-lag inde

In another test of effective (directional) connectivity, the similarity between weighted-phase-lag-index-derived networks during rest and CCEP networks was computed over a range of frequencies, with the best agreement occurring around 30 Hz (**Figure 3C**). While the lower bound of the uncorrected 95% was higher than zero at all frequencies tested, overall the agreement between weighted phase lag index connectivity and the CCEP network was poor, and the highest similarity score is 0.10 ± 0.15 std. Examining the individual values, the average weighted phase lag indices between the stimulation site and channels with above-threshold CCEPs were higher than between the stimulation site and channels with below-threshold responses, particularly in the 1-10 Hz and 25-35 Hz ranges, but the variability was also very high (**Figure 3D**).

### Structural Connectivity

#### Probabilistic tractography

Preoperative diffusion tensor imaging (DTI) was conducted in a subset of participants where the networks were calculated between the area around each stimulation site and the areas around the brain locations of the recording channels for a total of 34 stimulation sites across 8 participants. Across 34 stimulation sites, CCEP networks had a higher average similarity to structural networks than any of the functional networks tested, with tractography networks from the stimulation channel to the recording channel averaging a similarity score of 0.68 ± 0.21 std and tractography networks from the recording channel to the stimulation channel averaging a similarity score of 0.67 ± 0.23 std (**Figure 4A**). There was no statistically significant difference between the stimulation channel to the recording channel group and the recording channel to the stimulation channel (p = 0.51; t-test). Further, CCEP were more similar to structural networks than to correlation networks (paired t-test using the subset of participants with diffusion tensor imaging, p = 3.4e-09 and p = 3.6e-08, respectively). Channels with above-threshold CCEPs had an average normalized tract percentage of 0.41 ± 0.29 std from the stimulation channel to the recording channel and 0.40 ± 0.31 std from the recording channel to the stimulation channel, compared to 0.019 ± 0.072 std and 0.017 ± 0.069 in the remaining, below-threshold responsive channels (**Figure 4B**).

**Figure 4.**
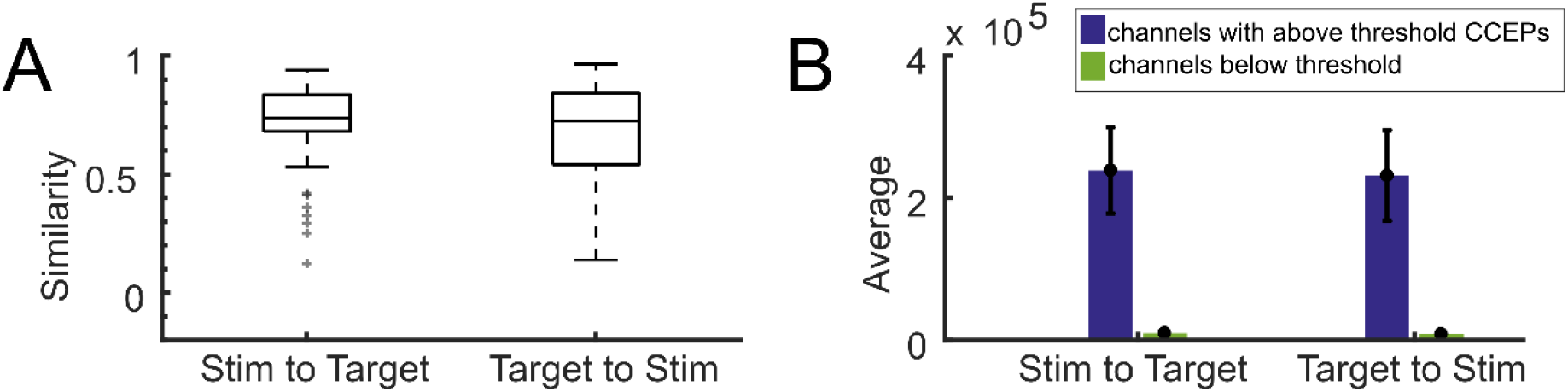
Structural connectivity shows high similarity to stimulation-evoked networks. **A**. Boxplots showing the medians and confidence bounds of the similarity scores diffusion tensor imaging networks between stimulation sites and recording sites in both directions and CCEP networks across 34 for different stimulation sites. Diffusion tensor imaging tractography is a directed measure of connectivity, so calculations from the stimulation site to the recording channel and from the recording channel to the stimulation site are both represented. **B**. The average unnormalized DTI connectivity streamline count for all channels with (blue) or without (green) an above-threshold CCEP. The presence of a CCEP is defined as any average evoked potential that reaches an absolute value at least 6 standard deviations above baseline mean.Uncorrected confidence intervals (black error bars) were determined by naive bootstrap resampling. Error bars reflect 95% confidence intervals.

### Controlling for Distance

All these estimates of connectivity share a common cofounding factor – distance from the recording channel to the stimulation channel. Correcting for distance using a partial correlation approach substantially lowered the similarity between CCEP networks and the other measures across the board (**Figure 5**). Still, all connectivity estimates except weighted phase lag indices maintained a significant similarity to CCEP networks even when corrected for distance. While uncorrected network similarity showed DTI as significantly more similar to CCEP networks than correlation, corrected similarity measures between these two methods were no longer significantly different (p = 0.30 for tractography from the stimulation channel and p = 0.19 for tractography to the stimulation channel).

**Figure 5:**
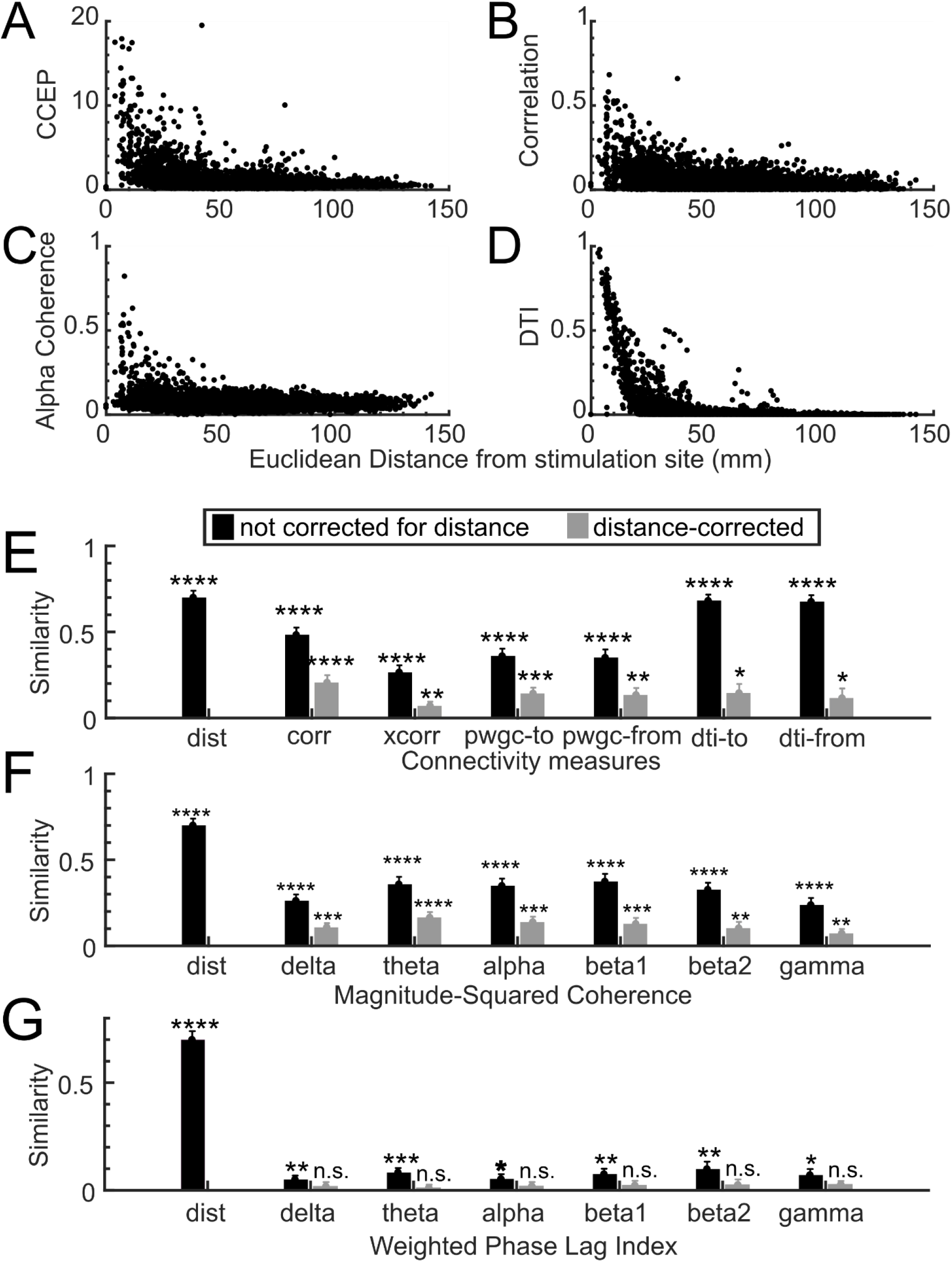
Network measures used in this paper share distance as a confounding factor. **A-D** show the relationship between a few selected measures and the Euclidian distance between the stimulation channel and the recording channel. **A**. A scatter plot of the normalized absolute peak in the 250 ms post-stimulation (CCEP) at the recording channel and distance from the stimulation channel. **B**. A scatter plot of the correlation of the recording channel and the stimulation channel and distance between those channels. **C**. A scatter plot of mean normalized magnitude coherence in the 8-12 Hz range between the recording and the stimulation channels and distance between those channels. **D**. A scatter plot of the normalized diffusion tensor imaging tractography strength and the Euclidean distance between the recording and stimulation channels. **E-G**. The average similarity between CCEP networks and the indicated connectivity measure (E: various metrics, F: magnitude-squared coherence across frequency bands, G: weighted phase lag index across frequency bands) without (black bars) or with (grey) distance correction across 34 stimulation sites. Error bars indicate the standard error of the mean. Method labels: distance (dist), correlation (corr), cross-correlation (xcorr), pairwise granger causality (pwgc), and diffusion tensor imaging tractography (dti). Frequency bands: 1-4 Hz (delta), 4-8 Hz (theta), 8-12 Hz (alpha), 15-20 Hz (beta1), 20-30 Hz (beta2), 30-50 Hz (gamma). Significance thresholds: p < 0.05 (*), p < 0.01 (**), p < 0.001 (***), p < 0.0001 (****).

Though the similarity scores decreased when correcting for distance, distance did not have a homogenous effect across connectivity metrics. Subdividing the recording channels into local (within 3cm of the stimulation site) and distant (further than 3cm from the stimulation site) groups revealed that functional and effective connectivity estimates during rest corresponded best with the CCEP networks at local channels, while structural connectivity metrics demonstrated similar relations to stimulation responses at both local and distant channels (**Figure 6**). The method with the highest similarity to the local CCEP networks was correlation, and the method with the highest similarity to the distant channel CCEP networks was DTI from the stimulation site to the recording site (**Figure 6**). Breaking the functional connectivity down into its frequency components with magnitude-squared coherence, the high beta and gamma frequency bands were not similar to distant channel CCEP networks, with mean similarity values close to zero.

**Figure 6:**
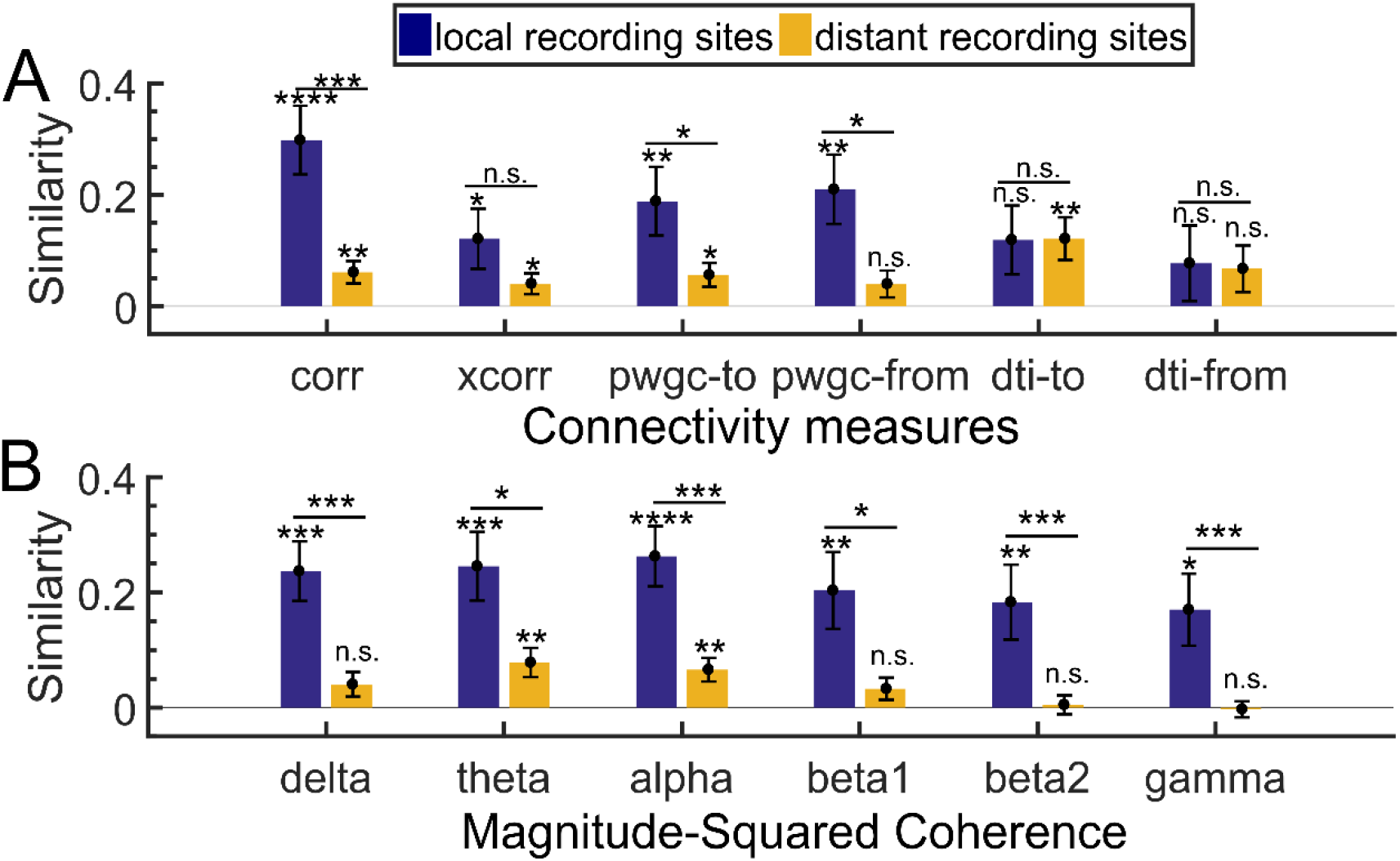
Network similarity for local and distant channels relative to the stimulation site. **A-B**. The mean distance-corrected similarity between CCEP networks and the indicated connectivity measure across 34 stimulation sites when channels are split by distance to the stimulation site into two groups: local (dark blue) or distant (orange). Local channels are within 3cm of the stimulation channel, as measured by Euclidean distance, and distant channels are outside that radius. Error bars indicate the standard error of the mean. Method labels: distance (dist), correlation (corr), cross-correlation (xcorr), pairwise granger causality (pwgc), and diffusion tensor imaging (dti). Frequency bands: 1-4 Hz (delta), 4-8 Hz (theta), 8-12 Hz (alpha), 15-20 Hz (beta1), 20-30 Hz (beta2), 30-50 Hz (gamma). Significance thresholds: p < 0.05 (*), p< 0.01 (**), p < 0.001 (***), p < 0.0001 (****).

## Discussion

We systematically compared CCEP networks to functional, effective, and structural connectivity networks. Comparisons where we did not control for distance suggest that structural connectivity derived through diffusion tractography was the strongest correlate of connectivity represented by the CCEP network. However, controlling for distance abolished the advantage of structural connectivity above functional and effective estimation methods. By segmenting channels into two groups – local and distant – we showed that, even controlling for distance, structural connectivity is the most similar to distant CCEP spread while local CCEP networks reflect functional connectivity measures.

Uncorrected functional connectivity metrics showed a reasonable similarity to CCEP connectivity. Of this group, cross-correlation showed the least similarity to CCEPs, even though cross-correlation has been used to demonstrate stable networks in EEG across time [25]. Unlike the assumption of simultaneity under correlation, cross-correlation is based on a more plausible biophysical assumption, that two connected regions may lag each other in activity. However, the inclusion of a variable lag within a relatively large window increased the noise of the estimation by raising the floor of possible values. Thus, cross-correlation found many connections between channels that did not respond to stimulation. Magnitude-squared coherence showed the highest similarity in spatial distribution to CCEPs in the 10-20 Hz range, consistent with the notion that lower frequency oscillations are more spatially distributed and more dependent on white matter connections than high frequency oscillations [36–38].

Despite categorization of stimulation as a measure of effective connectivity, the effective connectivity measures chosen here were overall less similar to CCEP networks than functional and structural connectivity measures. This conclusion was also found in another study comparing Granger causality, cross correlation, and the CCEP network [39]. Weighted phase lag index fared particularly poorly in our hands, though both measures tended to under-identify channels that would be responsive to stimulation. Though Granger causality is a directed measure of connectivity, direction seemed to matter little in corresponding to the CCEP network. This lack of difference could be because stimulation may activate white matter tracts in both orthodromic and antidromic directions, or it could be due to the high degree of reciprocal connections between cortical areas [4,16,40,41]. Indeed, stimulation location relative to the grey and white matter should be taken into account in future work, as it has been demonstrated that white matter stimulation induces more widespread network responses with depth electrodes [9].

A possible confound of this work and comparisons to other studies, however, is that all the data included here involves stimulation from sEEG, intraparenchymal electrodes in different regions of cortical and subcortical locations. While there has been work on the physiology of stimulation responses from intraparenychymal electrodes (particularly with respect to DBS for movement disorders), there still remain many questions about the relationship between brain responses to stimulation on the pial surface versus stimulation inside the cortex. Undoubtedly, the location of the stimulation has a substantial impact on which neural elements are modulated. Notably, much of the recent work involving characterizing CCEPs and their consistent waveform characteristics to date have been on CCEPs induced by stimulation via superficial grid electrodes, particularly in the investigation of involved neural networks [4,8,8,16] and as related to underlying white matter tracts and brain states [22]. However, this current study represents one of but a few to explicitly explore the relationship between stimulation of both cortical and subcortical structures and connectivity. This remains an important point as there is speculation that CCEPs are a representation of network interactions between cortical and subcortical pathways as opposed to a purely local or cortical evoked population response induced via white matter connections [42–44].

In estimates of similarity not controlling for distance, connectivity based on diffusion tractography imaging (DTI) was most similar to connectivity based on CCEPs. DTI is also a directed measure of connectivity, though in practice the directions differed very little from each other because diffusion happens in both orthodromic and antidromic directions along axons. Mechanistically, stimulation is thought to primarily excite axons [45,46], so the agreement between CCEP spread from electrical stimulation and structural connectivity was perhaps not surprising. Still, it is important to note that both CCEPs and DTI shared a common cofounding factor – distance from the stimulation site [46]. In addition, practical limitations of collecting data in the clinical setting resulted in heterogeneous DTI data collection parameters; higher quality data set may improve results.

After correcting for distance, the landscape changed dramatically. Weighted phase lag index was the only metric, of the ones we tested, to no longer be significantly similar to CCEP networks after distance correction. Coherence in the gamma band was relatively unaffected by distance corrections, which could be due to its relatively constrained spatial distribution in the first place. While estimates before correcting for distance had DTI most similar to CCEP networks, distance correction abolished this advantage. There was no statistically measurable difference in distance-corrected similarity between DTI, correlation, Granger causality, and coherence in all bands except delta and gamma. These results would seem to indicate that, once corrected for distance, many methods of estimating connectivity perform reasonably similarly in predicting stimulation response. This is perhaps why these various measures have been used to show that CCEP can reflect functional and effective connectivity in the brain in past studies [17,20,39].

However, overall comparable distance-corrected similarity scores did not mean that this similarity was homogenously applicable across the entire pool of channels across the brain. Segmenting the channels into two groups by distance to the stimulation site revealed that structural connectivity was more robust as one gets further away from the stimulation site, even after correcting for distance. On the other hand, correlation again rose to the top as being most similar to channels closest to the stimulation site. Structural connectivity may have failed to predict local responsive channels because of its poor spatial resolution. Though MRI as a modality has much higher spatial sampling than sEEG, diffusion tractography requires the creation of reasonably large regions of interest as seed and target masks (up to 1 cm in diameter), limiting its utility in estimating connectivity of overlapping ROIs. On the other hand, correlation and coherence, particularly in the higher frequencies, are known to drop off dramatically with distance in the cortex [47]. Our results affirm that different methods may be more appropriate within specific spatial regimes. In addition, as there is evidence that the CCEP network is different depending on the brain region [9,16,22], future work with more widespread and repeated sampling across brain regions could help resolve if there are regional differences in the similarities between these metrics and the CCEP network. Of course, an added dimension is comparison of these connectivity metrics during other conditions (and not just during resting state) such as a language task or other activities [39] could illuminate further whether these metrics and their similarity to the CCEP network map to state and state transitions as opposed to just brain region or distance [22].

Our results suggest that structural connectivity methods produce very similar networks to those derived by single pulse electrical stimulation. However, this similarity was driven in large part due to the underlying relationship between distance and connectivity. This relationship is likely due to a variety of factors, including volume conduction, the cumulative probability of terminating a tract, and the underlying tendency of brain regions to form both physical and functional connectivity with neighboring areas. Still, distance-corrected connectivity was more suited for the purposes of determining which non-invasive methods of connectivity estimation may help predict the effects of stimulation. Taken together, our results indicate that many methods may suffice, though functional and effective methods seemed particularly well-suited for predicting local stimulation effects and this relationship could be crucial when designing stimulation paradigms for clinical and research purposes.

## Data Availability

All data underlying the study will be made available upon reasonable request.

## Acknowledgments

We would like to especially thank the patients who participated for their time and help. We thank Erica Johnson, Gavin Belok, Kara Farnes, and Mia Borzello for technical assistance, particularly in the MRI reconstruction and registration, and Enterprise Research Infrastructure & Services at Partners Healthcare for their in-depth support and for the provision of the ERISOne Linux Computing Cluster. This work was supported by NINDS-K24 [K24-NS088568-01A1]; the Tiny Blue Dot Foundation; and the United States Department of Energy Computational Sciences Graduate Fellowship [DE-FG02-97ER25308] to BC. This research was sponsored by the U.S. Army Research Office and Defense Advanced Research Projects Agency (DARPA) under Cooperative Agreement Number W911NF-14-2-0045 issued by ARO contracting office in support of DARPA’s SUBNETS Program. The views, opinions, and/or findings expressed are those of the authors and should not be interpreted as representing the official views or policies of the Department of Defense or the U.S. Government.

